# Use of c-peptide as a measure of cephalic phase insulin release in humans

**DOI:** 10.1101/2022.05.03.22274582

**Authors:** Alexa J. Pullicin, Sean A. Newsom, Matthew M. Robinson, Juyun Lim

**Affiliations:** Department of Food Science and Technology, Oregon State University, Corvallis, OR 97331, USA; School of Biological and Population Health Sciences, Oregon State University, Corvallis, OR, United States

**Author notes:** Correspondence to:* Juyun Lim, Ph.D., Department of Food Science and Technology, Oregon State University, 100 Wiegand Hall, Corvallis, OR 97331, USA.

**Keywords:** cephalic phase, preabsorptive insulin release, c-peptide, oral stimulation, individual differences

## Abstract

Cephalic phase insulin release (CPIR) is a rapid pulse of insulin secreted within minutes of food-related sensory stimulation. Understanding the mechanisms underlying CPIR in humans has been hindered by its small observed effect size and high variability within and between studies. One contributing factor to these limitations may be the use of peripherally measured insulin as an indicator of secreted insulin, since a substantial portion of insulin is metabolized by the liver before delivery to peripheral circulation. Here, we investigated the use of c-peptide, which is co-secreted in equimolar amounts to insulin from pancreatic beta cells, as a proxy for insulin secretion during the cephalic phase period. Changes in insulin and c-peptide were monitored in 18 adults over two repeated sessions following oral stimulation with a sucrose-containing gelatin stimulus. We found that on average, insulin and c-peptide release followed a similar time course over the cephalic phase period, but that c-peptide showed a greater effect size. Importantly, when insulin and c-peptide concentrations were compared across sessions, we found that changes in c-peptide were significantly correlated at the 2 minute (*r* = 0.50, *p* = 0.03) and 4 minute (*r* = 0.65, *p* = 0.003) time points, as well as when individuals’ peak c-peptide concentrations were considered (*r* = 0.64, *p* = 0.004). In contrast, no significant correlations were observed for changes in insulin measured from the sessions (*r* = −0.06-0.35, *p* < 0.05). Herein, we detail the individual variability of insulin and c-peptide release during the cephalic phase period, and discuss why c-peptide may be a more appropriate metric to represent insulin secretion.

## 1. Introduction

Cephalic phase insulin release (CPIR) is a small, transient spike of insulin that aids in maintaining glucose homeostasis following nutrient ingestion (1–4). Unlike postprandial insulin release, whereby insulin concentration rises following nutrient accumulation in the gut, CPIR occurs within minutes of meal onset and relies on the activation of neural signals originating from sensory (e.g., gustatory, olfactory) inputs (5,6). Nonetheless, the underlying sensory and neural basis of the response has not been fully elucidated. Progress toward understanding such mechanisms has been hindered by a small observed effect size and high variability within and between studies in humans (7–10). Implementing study protocols that limit these issues will be key for progressing CPIR research.

A critical element of a CPIR study protocol is how the degree of insulin secretion is estimated. Within the current body of CPIR literature, nearly all studies use peripheral insulin concentrations to assess the degree of the response (7,10). However, peripheral insulin concentrations may not be the optimal metric to estimate pancreatic insulin secretion, i.e., the actual extent of CPIR. This is because approximately 40-80% of insulin is removed during first-pass transit through the liver prior to its delivery to peripheral circulation (11–14), meaning peripheral insulin is the net balance of insulin secretion and hepatic clearance. Consequently, peripheral insulin concentrations could underestimate insulin secretion by the pancreas, thereby hindering measurement of CPIR. Furthermore, given that the degree of insulin clearance differs significantly between individuals and under certain physiological circumstances (15,16), hepatic insulin clearance could enlarge the variability of CPIR.

C-peptide is a short polypeptide connecting the A and B chains of insulin within the proinsulin molecule (i.e., the insulin precursor), which is cleaved to yield bioactive insulin and c-peptide within the pancreatic beta cell. C-peptide is secreted from pancreatic beta cells in equimolar concentrations as insulin (17), but has negligible hepatic extraction (11) and a slower and more constant rate of degradation than insulin (18). Its half-life (20-30 minutes) is longer than that of insulin (3-5 minutes) and thus it circulates at concentrations approximately five times higher in the systemic circulation (19). For these reasons, peripheral c-peptide concentration has been used as a proxy of insulin secretion in a variety of fields and clinical situations (19–21). Likewise, using c-peptide as an index of CPIR could circumvent the limitations associated with peripheral insulin measurement.

The primary objective of this study was to evaluate the use of c-peptide as a suitable indicator of CPIR in humans. To achieve this goal, we examined changes in insulin and c-peptide concentrations over the course of the cephalic phase period (0-8 minutes) following exposure to a gelatin-based sucrose stimulus at two repeated sessions. We then compared changes in plasma insulin and c-peptide concentrations and evaluated their inter- and intra-individual variability. Finally, we determined whether changes in insulin and c-peptide concentrations correlated across the repeated sessions.

## 2. Participants and Methods

### 2.1. Participant Eligibility

Eighteen individuals (10 M, 8 F; mean ± SD = 27.2 ± 5.4 years) participated in the study. Eligibility criteria included 1) age between 18 and 35 years old, 2) self-reported healthy, 3) not a smoker, 4) not diabetic, 5) not pregnant, 6) no oral piercings, 7) no known food allergies, 8) no known taste or smell disorders, 9) not on prescription pain or beta-blocker medications, 10) no sickle cell disease, 11) no symptoms of a heart condition within 6 months, 12) on a relatively stable diet (i.e., meals and snacks typically consumed at similar times on weekdays), 13) no calorie-restricted diet, 14) on a normal weekday sleeping pattern (i.e., going to sleep at a similar time [± 1 hour] and waking up at a similar time [± 1 hour] on typical weekdays), 15) weight of > 50 kg, 16) no anxiety to blood draws, 17) body mass index (BMI) between 18.0 kg/m^2^ and 34.9 kg/m^2^, and 18) normal taste sensitivity to carbohydrates (see section 2.2.2).

Participants were asked to comply with the following restrictions prior to all visits: no consumption of foods or beverages except water for 1 hour (single screening visit) or 8 hours (two test visits), no dental work within 48 hours, no consumption of alcohol or use of cannabis products within 12 hours, no use of menthol products within 1 hour, and no physically demanding activity the morning of the visit. The study was conducted in accordance with the Declaration of Helsinki and was approved by the Oregon State University Institutional Review board and registered under the Clinical Trial registry (NCT02589353). All participants gave written informed consent and were paid for time in study.

### 2.2. Experimental Design and Procedures

#### 2.2.1. Online Screening

Potential participants were directed to a Qualtrics online eligibility survey. Individuals who met eligibility criteria 1-16 (see section 2.1 above) were invited to an in-person screening visit.

#### 2.2.2. Screening Visit

During the screening visit, body measurements were taken and taste sensitivity to carbohydrates (glucose and a maltooligosaccharide preparation) was evaluated. The outcomes were used to further screen participants for eligibility criteria 17-18.

##### Body measurements

Height and weight were measured using a beam scale with a height rod to the nearest 0.5 cm and 0.1 kg, respectively. Waist and hip circumferences were measured using a tape measure to the nearest 0.5 cm at the smallest point of the waist and widest portion of the hips, respectively (WHO, 2008). Participants were accepted if their BMI was between 18.0 kg/m^2^ and 34.9 kg/m^2^.

##### Taste sensitivity measurements

To confirm normal carbohydrate taste sensitivity, participants performed six sets of discrimination tests (triangle method) using three concentrations of glucose and a maltooligosaccharide preparation (see section 2.3.1 below). Carbohydrate stimuli and blanks were presented to participants by swabbing the sample across the dorsal tip of the tongue using a cotton-tipped plastic applicator. Participants were accepted if they correctly responded to at least 3 triangle tests.

#### 2.2.3. Test visits

Each participant attended two identical test visits. Upon arrival to the test visits, blood glucose was measured via a finger prick blood sample (CONTOUR® NEXT ONE, Ascensia Diabetes Care, Parsippany, NJ, USA). A blood glucose concentration of less than 110 mg/dL was accepted for the participant to continue with the session.

An intravenous cannula was inserted into a superficial vein in the antecubital space of the arm and secured for the duration of the session. Saline was used throughout the session to maintain patency of the line. Following cannulation, the lower forearm and hand of the cannulated arm was wrapped in a heating pad to allow for collection of arterialized venous blood samples, which more closely represents metabolite levels in arteries (22). Participants rested for 15 minutes after cannulation to allow metabolite levels to stabilize. After the resting period, three baseline blood samples were collected at 5-minute intervals (−15, −10, and −5 minutes prior to oral stimulation; see **Fig. 1** for blood collection timeline). Each collection involved removal of a waste sample (infused with saline) followed by a 3 mL blood collection. At the time of oral stimulation (0 minutes), participants placed the entire gelatin stimulus (see section 2.3.2 below) in their mouth and chewed for 45 seconds. Participants were instructed the chew the sample in the same manner that they would normally chew a bite of food. After 45 seconds had elapsed, participants expectorated the stimulus. Blood samples (3 mL) were then collected at 2, 4, 6, and 8-minute time points following the start of oral stimulation.

**Fig. 1.**
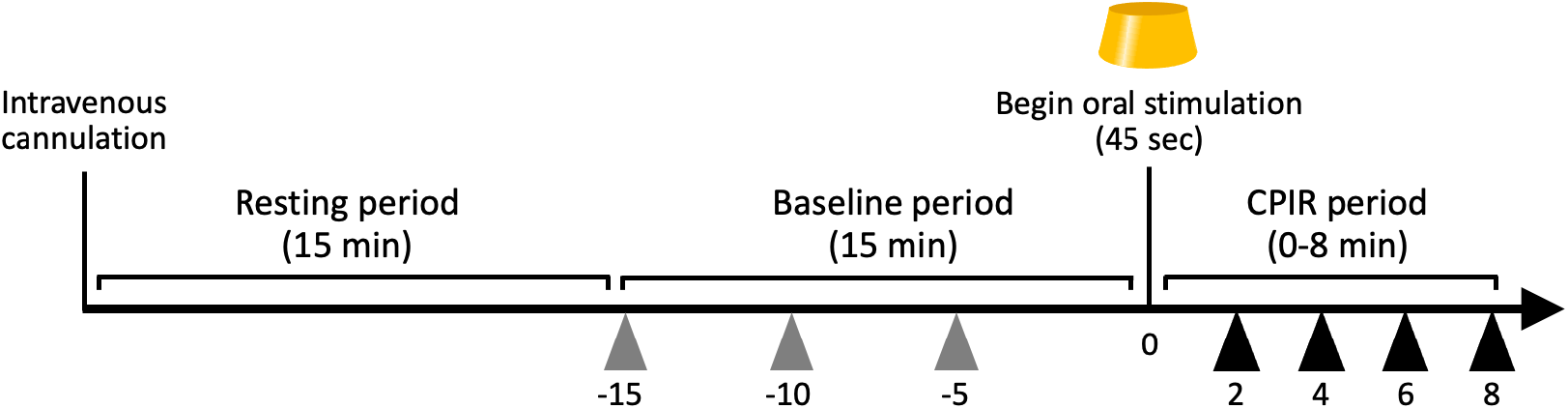
Overview of blood draw protocol. Participants received the model gelatin stimulus at the 0 minute time point and performed modified sham-feeding for 45 seconds. Arrows labeled with numbers represent blood collection time points (in minutes) during the baseline (gray) and CPIR (black) periods. Each participant underwent this protocol on two separate days.

### 2.3. Stimuli

#### 2.3.1. Stimuli for taste sensitivity

Glucose and a maltooligosaccharide preparation were used to measure carbohydrate taste sensitivity. The profile of the maltooligosaccharide preparation is described in Balto et al. ((23); see 90EI/70ES-CSS). Stimuli were prepared in ultrapure water at 56, 100, and 180 mM at least 12 hours before the session in which they were used to allow anomeric equilibration of reducing sugars (24). Stimuli and blanks were prepared with 5 mM acarbose to inhibit oral hydrolysis of the maltooligosaccharide preparation by salivary α-amylase. Taste stimuli were brought to room temperature (20-22 °C) before the testing session.

#### 2.3.2. Oral stimulus to elicit CPIR

A 15-cc gelatin-based model stimulus was used to elicit CPIR. A solid stimulus was chosen for this study over a solution because solid stimuli require more oral manipulation, which in turn produces greater oral stimulation and could elicit a greater CPIR (8,25). The stimulus was prepared with deionized water and consisted of 34% w/v (1M) sucrose and 11% w/v gelatin. Yellow food coloring was added to simulate the sensory experience of eating a similar food item. The gelatin disks (in the shape of a conical frustum) were approximately 1.9 cm tall, and 2.8 and 3.4 cm at the smallest and widest parts across, respectively. The stimulus was brought to room temperature (20-22 °C) before serving.

### 2.4. Biochemical Analyses and Specimen Processing

#### 2.4.1. Blood specimen processing

Blood samples were transferred to heparinized vacuum tubes as they were collected and stored on ice. Following the testing session, blood samples were centrifuged at 1000 x g for 10 minutes and the plasma was aliquoted and frozen in a clinic freezer prior to transporting to long-term storage at −80 °C.

#### 2.4.2. Biochemical analyses

Plasma insulin and c-peptide were measured using the Insulin ELISA and C-peptide ELISA kits from Alpco (Salem, NH, USA) without modification. To measure plasma glucose, 3 μL of plasma samples were added to wells of a microtiter plate followed by 300 μL of Infinity(tm) Glucose Hexokinase Liquid Stable Reagent (Thermo Scientific Inc, Middletown, VA, USA). The plate was incubated at 37 °C for 3 minutes then absorbance was read at 340 nm. Glucose concentrations were determined against a standard curve. Plasma samples were analyzed in duplicate for the three assays and repeated control samples were used across different plates.

### 2.5. Statistical Analyses

#### 2.5.1. Data handling

Fasting insulin, c-peptide, and glucose values for each participant were obtained by averaging their concentrations across −15, −10, and −5 minutes. To determine changes in insulin (Δ insulin), c-peptide (Δ c-peptide), and glucose (Δ glucose), the averaged fasting concentrations were subtracted from each post-stimulus time point value (i.e., concentration at 2, 4, 6, and 8 minutes) for each participant.

#### 2.5.2. Data analyses

Mean and standard deviation were used to describe participant age, anthropometrics, and fasting concentrations of plasma insulin, c-peptide, and glucose. The following statistical analyses were conducted:

1. Two-sample t-tests were performed to determine whether there were significant differences in age or anthropometric measurements between males and females.
2. To determine if Δ insulin, Δ c-peptide, and Δ glucose concentrations increased from baseline, one-sample t-tests were performed separately for each post-stimulus time point.
3. To determine if Δ insulin and Δ c-peptide values correlated within a sample, Pearson product-moment correlations were computed. Data from 2 and 4 minutes were considered since CPIR is generally reported to peak around 2 to 4 minutes following the onset of sensory stimulation. The same analysis was conducted using each participants’ peak Δ insulin and Δ c-peptide concentrations within 2 to 8 minutes, given the expected variability in the time course of insulin and c-peptide profiles within individuals.
4. To evaluate the consistency of insulin and c-peptide measurements across repeated sessions, the Pearson product-moment correlations were calculated for Δ insulin and Δ c-peptide concentrations obtained from the test sessions at 2 minutes and 4 minutes, as well as the peak (maximum) Δ values from each session.

Analyses were performed in Statistica 13 (StatSoft, Inc.), and the statistical significance criterion was set at *p* < 0.05.

## 3. Results

### 3.1. Participant Characteristics

Participant characteristics, including fasting (baseline) concentrations of insulin, c-peptide, and glucose are shown in **Table 1**. When data were grouped by sex, males had a greater mean waist circumference and waist-to-hip ratio than females. Conversely, BMI, hip circumference, and fasting concentrations of glucose, insulin, and c-peptide did not significantly differ between sexes. Furthermore, fasting concentrations of insulin, c-peptide, and glucose did not significantly differ between the two sessions within any grouping (i.e., male, female, all).

**Table 1.**
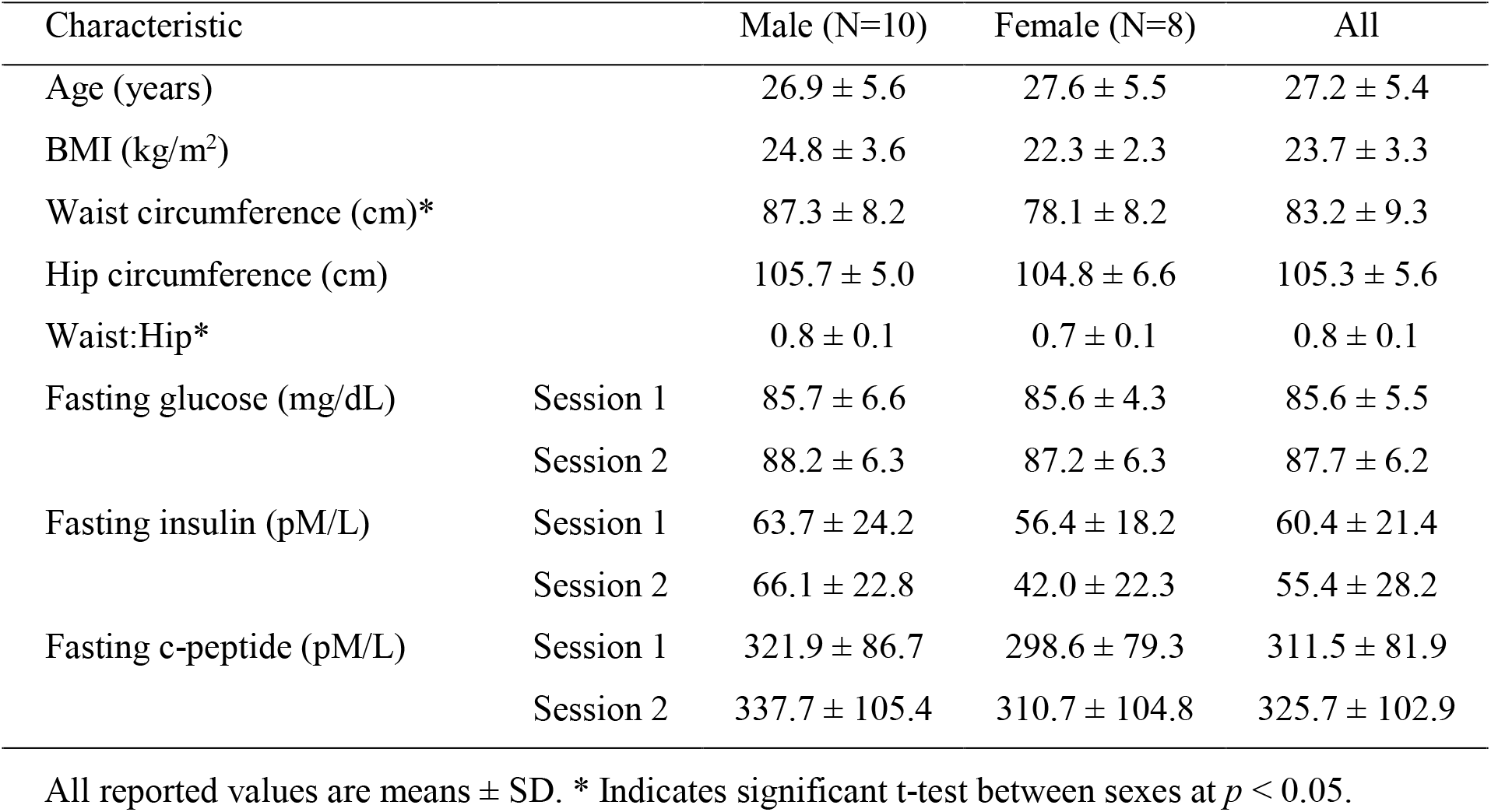
Characteristics of participants

| Characteristic |  | Male (N=10) | Female (N=8) | All |
| --- | --- | --- | --- | --- |
| Age (years) |  | 26.9 ± 5.6 | 27.6 ± 5.5 | 27.2 ± 5.4 |
| BMI (kg/m<sup>2</sup>) |  | 24.8 ± 3.6 | 22.3 ± 2.3 | 23.7 ± 3.3 |
| Waist circumference (cm)* |  | 87.3 ± 8.2 | 78.1 ± 8.2 | 83.2 ± 9.3 |
| Hip circumference (cm) |  | 105.7 ± 5.0 | 104.8 ± 6.6 | 105.3 ± 5.6 |
| Waist:Hip* |  | 0.8 ± 0.1 | 0.7 ± 0.1 | 0.8 ± 0.1 |
| Fasting glucose (mg/dL) | Session 1 | 85.7 ± 6.6 | 85.6 ± 4.3 | 85.6 ± 5.5 |
|  | Session 2 | 88.2 ± 6.3 | 87.2 ± 6.3 | 87.7 ± 6.2 |
| Fasting insulin (pM/L) | Session 1 | 63.7 ± 24.2 | 56.4 ± 18.2 | 60.4 ± 21.4 |
|  | Session 2 | 66.1 ± 22.8 | 42.0 ± 22.3 | 55.4 ± 28.2 |
| Fasting c-peptide (pM/L) | Session 1 | 321.9 ± 86.7 | 298.6 ± 79.3 | 311.5 ± 81.9 |
|  | Session 2 | 337.7 ± 105.4 | 310.7 ± 104.8 | 325.7 ± 102.9 |

All reported values are means ± SD. \* Indicates significant t-test between sexes at $p < 0.05$ .

### 3.2. Insulin vs. c-peptide

On average across 18 participants, there was a significant increase in both insulin and c-peptide concentration within 2 to 4 minutes after the onset of oral stimulation (see **Fig 2A and 2B)**. To confirm that the increase in insulin and c-peptide concentrations did not reflect a corresponding increase in glucose concentration, glucose was analyzed in plasma samples collected between 2 and 8 minutes. As expected, no significant rise in glucose (*p* > 0.05) was observed during the cephalic phase period (**Fig. 2C**). This suggests that the modified sham-feeding protocol did not allow a measurable amount of glucose to be absorbed. A similar trend for Δ insulin, Δ c-peptide and Δ glucose concentrations was observed at session 2 (data not shown). Furthermore, sex had no impact on Δ insulin or Δ c-peptide concentrations at any time point, or on peak Δ insulin and Δ c-peptide concentrations at either session (data not shown).

**Figure 2.**
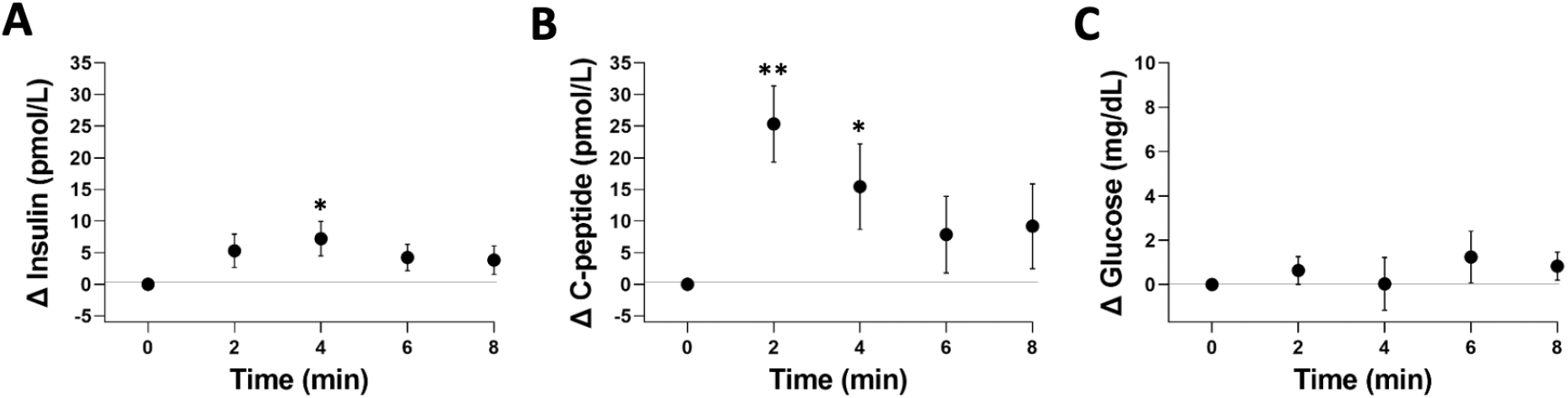
Change in insulin (A), c-peptide (B), and glucose (C) concentrations from baseline over the cephalic phase period observed at session 1. Values are group mean ± SEM. *,** indicate a statistically significant difference from baseline at *p* < 0.05 and 0.01, respectively. The zero-time point represents the group mean of the three baseline concentrations (−15, −10, and −5 minutes) and is set to 0 for reference.

When individual data were considered, the time to reach peak Δ insulin and Δ c-peptide concentrations varied between participants (see **Fig. 3**, filled black circles) as well as across sessions. At both sessions, most participants reached peak Δ insulin and Δ c-peptide concentrations at either 2 or 4 minutes (i.e., 78-83% for Δ insulin and 61-78% for Δ c-peptide; **Fig. 3**). Notably, we observed that only a small proportion of participants showed peak values at consistent times (e.g., at 2 minutes) across sessions; only 6 of 18 participants (33%) for Δ insulin and 4 of 18 participants (22%) for Δ c-peptide. This finding suggests that using a single time point (e.g., 2 or 4 minutes) to make conclusions about these responses could be misleading. Furthermore, peak Δ insulin and Δ c-peptide concentrations showed high variability across the participants: individuals’ peak concentrations ranged from −2 to 40 pmol/L for Δ insulin and from −8 to 80 pmol/L for Δ c-peptide (see **Fig. 3**, Peak).

**Figure 3.**
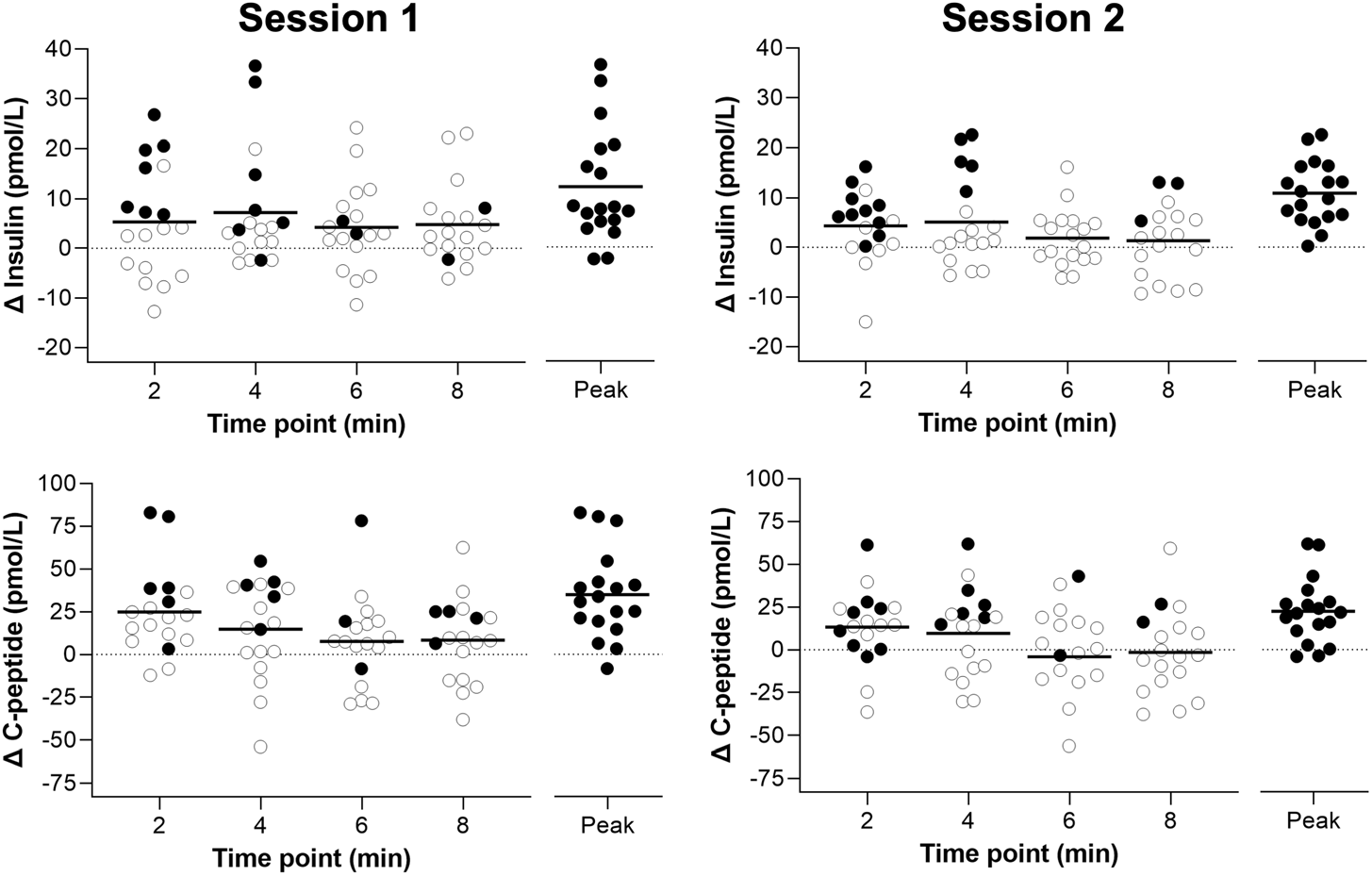
Change in insulin (top graphs) and c-peptide (bottom graphs) concentrations from baseline at session 1 (left panel) and session 2 (right panel). The circles at each time point represent the Δ insulin or Δ c-peptide concentration reached for each participant (N=18). Black circles indicate that the concentration was the participant’s peak concentration reached for the marker/session. These personal peak points are grouped together on the right of their corresponding graph (labeled “Peak” on the *x*-axis). A total of 18 circles are shown at each time point (2, 4, 6, 8 min) and the “peak” on the right. Solid black bars at each time point and the peak represent the mean of the grouping. The dotted line at zero represents the mean of the three baseline concentrations (−15, −10, and −5 minutes).

Next, we investigated the relationship between Δ c-peptide and Δ insulin concentrations. Correlation tests were conducted between Δ c-peptide and Δ insulin concentrations at 2 minutes and 4 minutes, since this is where a majority of participants reached their peak concentration. Correlations across the two sessions revealed only one significant relationship for the 2-minute time point of session 2, which was negative (*r* = −0.49, *p* = 0.04) (**Fig. 4**, left bottom). When peak values were considered, a positive significant relationship was found for only one of the two sessions (*r* = 0.55, *p* = 0.02). These results suggest that we cannot assume a direct relationship between Δ insulin and Δ c-peptide concentrations during the cephalic phase period.

**Figure 4.**
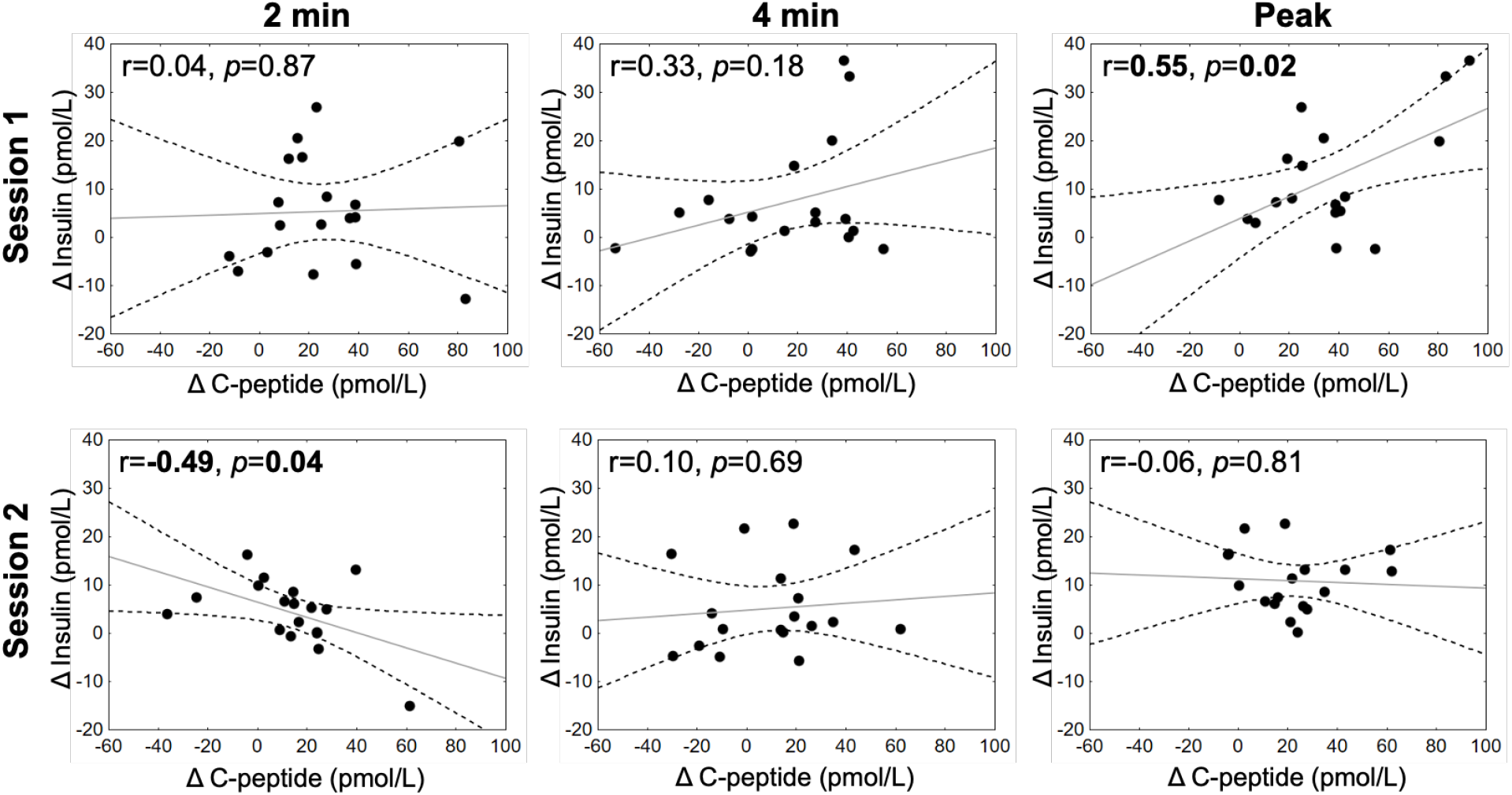
Correlations between Δ c-peptide (*x*-axis) and Δ insulin (*y*-axis) concentrations at 2 minutes (left panel), 4 minutes (center panel), and peak concentration measured within 2 to 8 minutes (right panel). The dashed lines represent the 95% confidence interval of the regression line (solid line). Correlation coefficients (*r*) and corresponding *p* values are indicated on the top left of each graph. Significant coefficients and p-values are bolded.

### 3.3. Consistency of insulin and c-peptide responses across repeated sessions

Pearson’s correlation coefficients were tested to determine whether Δ insulin and Δ c-peptide concentrations were consistent across repeated sessions (**Fig. 5**). We found that Δ insulin concentrations did not significantly correlate between the two repeated sessions at 2 minutes (*r* = 0.35, *p* = 0.15), 4 minutes (*r* = 0.04, *p* = 0.87), or when individuals’ peak values were considered (*r* = 0.06, *p* = 0.81). Conversely, Δ c-peptide concentrations significantly correlated between the sessions at both the 2-minute (*r* = 0.50, *p* = 0.03) and 4-minute time points (*r* = 0.65, *p* = 0.003), and when individuals’ peak values were considered (*r* = 0.64, *p* = 0.004).

**Figure 5.**
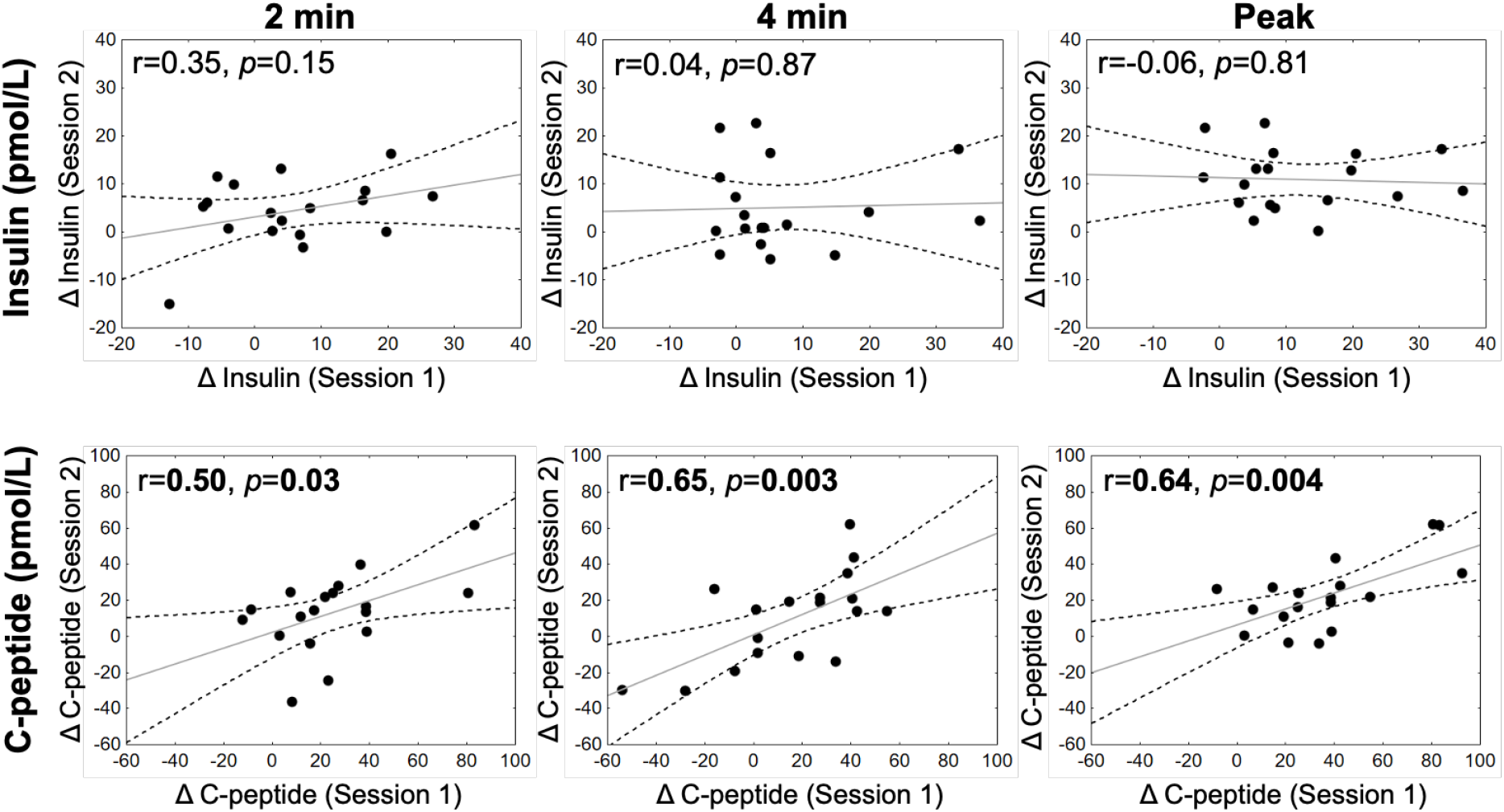
Correlations between Δ insulin concentrations (top graphs) and Δ c-peptide concentrations (bottom graphs) from two sessions (*x*-axis: session 1, *y*-axis: session 2). The figure depicts observations at 2 minutes (left panel), 4 minutes (center panel), and peak concentration measured within 2 to 8 minutes (right panel). The dashed lines represent the 95% confidence interval of the regression line (solid line). Correlation coefficients (*r*) and corresponding *p* values are indicated on the top left of each graph. Significant coefficients and p-values are bolded.

## 4. Discussion

### 4.1. Insulin and c-peptide profiles over the cephalic phase period

The present results show that on average, insulin and c-peptide release follow a similar time course during the cephalic phase period. Both markers reached their maximum concentrations within 2 to 4 minutes after oral stimulation began and subsided by 8 minutes (**Fig. 2A** and **2B**). A similar pattern of insulin release has been reported in other human CPIR studies, especially when no ingestion takes place (e.g., (25)). In contrast to insulin, few studies have measured c-peptide within the context of CPIR. Studies that have measured c-peptide show that, like we observed here, it follows a similar time course to that of insulin release over the cephalic phase period (26–29).

It should be recognized that the average reported peak time of CPIR varies across reports. For example, insulin has been reported to peak at 1 minute (30), 2 minutes (25), 3 minutes (31,32), 4 minutes (26,29,33), 5 minutes (27,28,34), or later (35,36) following the onset of sensory stimulation. The variability across these studies could be partially explained by differences in study protocols, including the duration of sensory stimulation (e.g., range from <1 minute to 20 minutes or greater) and blood sampling procedure (e.g., sampling every 1, 2, or 5 minutes). However, we observed differences in peak times even when all participants followed the same protocol, indicating that there are differences in the time course of cephalic phase responses across individuals (see section 4.2 below). These individual differences are likely to contribute to the variability reported across studies as well.

While the averaged time courses of Δ insulin and Δ c-peptide were generally similar, the present data show a significant difference in their magnitudes. The mean change in c-peptide concentration (range of ∼7 to 25 pmol/L; **Fig. 2B**) was typically 2 to 5 times greater at each measured time point compared to that of insulin (range of ∼4 to 7 pmol/L; **Fig. 2A**), except for Δ c-peptide concentrations at 6 and 8 minutes of session 2, which were lower than baseline. When participants’ Δ peak concentrations were considered (**Fig. 3**), a similar trend of higher c-peptide (session 1 mean = 35 pmol/L, session 2 mean = 23 pmol/L) versus insulin (session 1 mean = 12 pmol/L, session 2 mean = 11 pmol/L) was observed. The greater magnitude of Δ c-peptide observed during the cephalic phase period is presumably due to its limited hepatic degradation (cf. c-peptide is mostly extracted by the kidneys; (11)). Accordingly, this has implications for how the degree of CPIR is estimated when using blood sampled from peripheral circulation. Other studies using comparable stimuli (i.e., a model sucrose stimulus), wherein participants performed modified sham-feeding (25,28,34), reported Δ insulin concentrations that were similar to those observed here (approximately 3 to 19 pmol/L across time points within the cephalic phase period).

Only one study has measured c-peptide in the context of CPIR while using a comparable model stimulus to that used here along with a modified sham-feeding protocol (28). While that study found a small but significant increase in insulin concentration (up to ∼3 pmol/L) during the cephalic phase period following mouth rinsing with sucrose solution, the increase in c-peptide (up to ∼6 pmol/L) did not reach statistical significance due to large error terms observed. Nonetheless, other studies measuring c-peptide have reported a significant increase during the cephalic phase period (26,27,29). Teff and others (26) conducted experiments wherein participants were tested under both modified sham-feeding and ingestion protocols using a complex food (peanut butter sandwich). They reported, under both protocols, a significant increase in c-peptide concentration that was about 2.5 times greater than the increase in insulin at its maximum point. Two other studies (27,29) that involved ingesting complex food items (i.e., muffin, pizza) also reported increases in c-peptide concentration about 5 times higher than that of insulin at their maximum points. The current findings strongly support previous reports that c-peptide shows a similar time course of release as insulin during the cephalic phase period, but generally increases by a greater magnitude.

### 4.2. Individual differences in insulin and c-peptide responses during the cephalic phase period

This study shows for the first time in detail that there are substantial individual differences in the time course and the magnitude of insulin and c-peptide responses during the cephalic phase period. Across the two repeated sessions, most participants reached peak Δ insulin or Δ c-peptide concentrations at either the 2 or 4 minute time point (61-83% across the two markers and sessions, shown as filled black circles in **Fig. 3**). Nonetheless, a handful of individuals reached peak Δ concentrations at 6 or 8 minutes for insulin (17-22% of participants across sessions) and c-peptide (17-39% of participants across sessions). Importantly, the time at which participants reached their peak Δ concentration was not necessarily consistent between markers or sessions. In addition, we found that the magnitude of insulin and c-peptide responses vary considerably. As shown in **Figure 3**, peak Δ concentrations ranged from −2 pM to 40 pM for insulin and −8 pM to 80 pM for c-peptide. Note, however, that both markers showed a similar degree of variability across the group of participants when differences in scales were taken into consideration.

Individual differences in CPIR time course and magnitude are rarely detailed in human literature. Studies that comment on individual differences generally report that measured insulin variability is high across (37–40) and within (37,39) individuals. Recognizing these differences in CPIR, some authors have proposed dividing participants into “responder” and “non-responder” groups (25,38,41), a concept suggesting that some individuals produce measurable CPIR to a specific stimulus under the test conditions while others do not. However, this division could have limitations, particularly when measuring insulin. For instance, as shown in the present study, there is considerable variability even within a single individual across repeated sessions (see **Fig. 5**, top panel). In an effort to determine potential sources of this variability, some authors have investigated the role of specific factors on CPIR magnitude. Factors tested thus far include BMI (30,40,42), salivary amylase activity (35), degree of dietary restraint (43,44), and glucose tolerance (27). Within the present study, we did not find any significant relationship between Δ insulin or Δ c-peptide concentrations and any factor we measured (i.e., age, anthropometrics; data not shown). This, however, could be due to relative homogeneity of age (18-35 years) and anthropometrics (e.g., a BMI mean ± SD of 23.7 ± 3.3) of the study participants. Definitive factors that play a role in CPIR variability have yet to be identified (see discussion in (8)).

To consider potential reasons for the individual differences in cephalic phase insulin and c-peptide release reported here, it is helpful to understand how insulin and c-peptide are secreted and cleared. First, it is well documented that there are individual differences in beta cell function (45,46), which contribute to differences in the degree of insulin and c-peptide secretion (recall that insulin and c-peptide are co-secreted in equimolar amounts). While this difference is most pronounced in conditions such as metabolic syndrome and type 2 diabetes (47), individual differences in the secretory capacity of the beta cell in healthy individuals have been reported as well (48,49). Another important consideration is that at least 70% of total secreted insulin and c-peptide is released in a pulsatile manner (50) with a typical period of about 5 minutes (51,52). Consequently, natural fluctuations in the concentration of insulin and c-peptide will exist between measured time points independent of CPIR following sensory stimulation. Next, some variability, particularly regarding insulin, could be due to differences in hepatic insulin clearance. About 40-80% of secreted insulin is cleared from the bloodstream during its first pass through the liver (14). The level of clearance differs across individuals, as well as on a minute-by-minute basis within an individual (14). Moreover, insulin undergoes additional clearance (albeit to a lesser degree) during glucose uptake at peripheral sites (e.g., adipose and muscle tissues) and at the kidney, the extent of which varies under different physiological circumstances (11,53). Note that the protocol of the present study largely bypasses this latter concern by measuring arterialized blood.

Together, the aforementioned factors can explain some of the differences observed regarding time course and magnitude of CPIR across individuals. The latter contributions in particular (i.e., insulin clearance and differences in insulin and c-peptide metabolism) could also explain our findings that Δ insulin and Δ c-peptide concentrations did not directly correlate with one another (see **Fig. 4**), indicating the two markers do not increase proportionally in peripheral circulation during the cephalic phase period. These differences should be considered when reporting data and making conclusions about the responses, since basing conclusions solely on averaged data may misrepresent findings. For example, averaging participants’ concentrations at each blood sampling time point can blunt the apparent magnitude of the response if individuals reach peak concentrations at different times. This variability could in return underestimate the effect sizes and mislead the overall conclusion that can be drawn.

### 4.3. Measure of c-peptide is more replicable than insulin across repeated sessions

One of the most surprising findings of this study was that Δ insulin concentrations at 2 minutes, 4 minutes, and the participants’ peak time did not correlate between repeated sessions (*r* = 0.04-0.35, *p* > 0.15; see **Fig. 5**). This could be due in part to differences in insulin clearance, as discussed above (see 4.2). In contrast, a significant correlation was found for Δ c-peptide concentrations at 2 minutes (*r* = 0.50, *p* = 0.03), 4 minutes (*r* = 0.65, *p* = 0.003), and when individuals’ peak values were considered (*r* = 0.64, *p* = 0.004). Together, these results suggest that the increase in c-peptide during the cephalic phase period is more replicable across repeated sessions than insulin.

While no other studies to date have specifically examined the replicability of c-peptide responses in the context of CPIR, four studies have examined the replicability of insulin responses across repeated sessions (33,37,39,40). Two of these studies reported that the CPIR measured was not replicable between sessions (37,39) while two other studies reported otherwise (33,40). Teff and others (33) tested normal weight men (N=15) under three repeated trials following ingestion of a flavored mousse dessert. The authors state that the reliability of the responses on the three days was highly significant (*r* = 0.82, *p* <0.001), although details on how they reached this conclusion (i.e., data analysis) was not explicitly stated. Simon and others (40) also reported that reproducibility was established by comparing both Δ insulin concentration and insulin area under the curve. These outcomes are intriguing given that they reached this conclusion after testing five normal weight participants following visual and olfactory presentation of a meal. The present findings suggest that measure of c-peptide may offer more reliable outcomes of CPIR across repeated sessions.

## 5. Conclusions

The primary objective of this study was to evaluate the use of c-peptide as an indicator of CPIR in humans. C-peptide is secreted in equimolar amounts to insulin but undergoes minimal hepatic extraction. This provides a firm basis for the use of peripheral c-peptide concentrations as an estimate of beta cell activity. Indeed, c-peptide showed a cephalic phase response with a similar average time course to that of insulin, albeit having a greater measurable effect size than insulin. In addition, significant correlations of Δ c-peptide concentrations across sessions, but not Δ insulin concentrations, indicate that c-peptide may be a more replicable measure of CPIR than peripheral insulin. Together, these findings demonstrate that c-peptide could be used as a reliable proxy of insulin secretion during the cephalic phase period. Finally, our results show that there are substantial individual differences in the time course and the magnitude of insulin and c-peptide responses during the cephalic phase period. This variability needs to be considered in future studies when reporting and making conclusions about the response.

## Data Availability

All data produced in the present work are contained in the manuscript

## Acknowledgements

We would like to thank Sandra Uesugi for her assistance with blood collection. AJP is an ARCS (Achievement Rewards for College Scientists) Scholar.

